# “An exploratory Integrated Moving Average Time Series Model of the initial outbreak of COVID-19 in six (6) significantly impacted Countries”

**DOI:** 10.1101/2020.07.09.20150136

**Authors:** Joseph E. Pascarella, Elaina Pascarella

**Affiliations:** Saint Joseph’s College, New York; University of Virginia

**Keywords:** COVID-19 Predictive Modeling, Moving Average Time Series, Consistent Deceleration Model

## Abstract

The 2019 Novel Coronavirus SARS-CoV-2 (COVID-19) is a single-stranded RNA virus that has threatened the lives of humans all over the globe. Government officials, policy makers and public health officials have been scrambling and struggling to “flatten the curve” to decelerate the prevalence and spread of COVID-19 given the significant economic destruction of the spread of the virus. Most “flatten the curve” models are based on Compartmental Models. This preliminary research is based on six (6) selected countries significantly impacted by COVID-19 and endeavors to build a new model based on moving averages lagged at different time periods to better hone in on the time the COVID-19 begins to decelerate using the date of first reported case and date of first reported death. This new model, the Consistent Deceleration Model (CDM) is based on each individual country’s date of Peak Increase in Mortality Rate (PINC MR) and the Moving Average since the peak increase in mortality rate (MA POSTINC). The CDM can be utilized of one of many quantitative tools to determine the strength of the deceleration of an infectious outbreak.

## Introduction

The 2019 Novel Coronavirus SARS-CoV-2 (COVID-19) is a single-stranded RNA virus that has threatened the lives of humans all over the globe (Cascella, Rajnik, Cuomo, Dulebohn & Napoli, 2020). The new virus was confirmed a pandemic by the World Health Organization (WHO) on March 11th 2020 and the primary symptom is a lower respiratory illness (WHO, 2020). The initial epidemiological investigations aimed the origin of COVID-19 in Wuhan, China in December of 2019 and was officially reported to WHO on December 31, 2019 (World Health Organization, 2020). The likely source of the virus is estimated at a 96% probability to originate from bats with a potential but still unknown intermediate host between bats and humans potentially being the pangolin (Andersen, Rambaut, Lipkin, Holmes, & Garry, 2020). The virus is similar to previous viral outbreaks such as the 2012 to present Middle East Respiratory Syndrome Coronavirus (MERS-CoV) that has a mortality rate of 35% and the 2002-2003 Severe Acute Respiratory Syndrome Coronavirus (SARS-CoV) that had a mortality rate of 10% (Wu & McGoogan, 2020).

COVID-19 is spread from human-to-human, and less understood from human-to-animal/animal-to-human via close contact, via direct contact with infected aerosols or touching fomite that is why the practice of “social distancing” has been implemented in countries across the world (CDC, 2020). While the virus has an incubation period of two (2) to fourteen (14) days with a median of five (5) days for symptoms to show, the virus is unprecedented in its asymptomatic group of carriers increasing unintentional spreading and proving difficult to track both prevalence and incidence (Gandhi, Yokoe, & Havlir, 2020).

The first confirmed case in the United States was reported to have occurred on January 21st 2020 in Washington State when a 30 year old man returned from a trip to China who initially had no symptoms until a fever the day of his hospital admission on January 16th 2020 (Holshue et al., 2020) and according to the CDC, the first COVID-19 death in the United States occurred on February 29th 2019 in Washington State (CDC, 2020). However, Santa Clara County in California was the first confirmed community infected with COVID-19, and postmortem testing from February 6th through the 17th confirmed earlier COVID-19 infections (Soucheray, 2020). The first reported case in China was on December 8th, 2020 (Wu & McGoogan, 2020) and France reported the first COVID-19 case on January 24th, 2020, Germany on January 27th, Italy on January 31st, and Spain on February 1st of 2020 (Johns Hopkins, 2020). Italy, Spain, France, and Germany surges in COVID-19 cases contributing to European data reporting overall deaths to have increased 20-30% this year (Wu, McCann, Kat, & Peltier, 2020).

### Pandemics and the Analytical Framework for “Flattening the Curve”

Politicians, policy makers and pubic safety officials have primarily deferred to public health officials to “Flatten the Curve” to mitigate the spread of COVID-19 and lessen the burden on health care infrastructure (Branas, 2020) and prevent further spread and prevalence of COVID-19. Flattening the curve is based on an epidemiological mathematical family of Compartmental Models first discussed by Kermack, McKendrick and Walker (1927) and Kermack and McKendrick (1933) and factors in the spread of disease and applied to epidemics using time as a variable (Cooke, 1979). These models have evolved to include other variables and vary in degree of statistical sophistication (Beretta, & Takeuchi, 1995) with the primary variable as time and rate of infection, and using prior rates of infection over time to predict future exposure and prevalence. The essential objective is to measure the spread over time and degree of prevalence for the purpose of predicting the intensity of the increase in rate and if the increase will continue in intensity or began to slow. This is critical for a number of reasons to ensure the public safety and a healthy environment.

## Methodology

The objective of this current exploratory analysis is to determine the moving average change in mortality rate in three (3) day intervals of COVID-19 from initial onset of the epidemic through peak increase while accounting for the changes over time. This analysis is specific to six countries (the most consistent top five in terms of reported overall deaths from COVID-19) and China, the country where the Index Case of COVID-19 was first reported (World Health Organization, 2020). The top five (5) countries include the United States (US), Italy, Germany, France, and Spain. This present analytical framework is an Integrated Moving Average Model (IMAM) using time, and the preceding rate of mortality (MR) to determine when the COVID-19 first displayed evidence of slowing in countries that remained in the top five (5) for overall number of deaths attributed to COVID-19 and an added measure, the moving average after the largest increase in change to predict the deceleration of the decrease.

The time series spans from March 15, 2020 (three days after COVID-19 was declared a pandemic) through April 30, 2020. The time series data points were three (3) day intervals from totaling seventeen (17) data points and sixteen (16) dynamic changes. The time series analytical framework used was an Integrated Moving Average Model (IMAM) that tested the moving average of the rate of mortality over time since the date the COVID-19 was declared a pandemic by the World Health Organization (WHO) on March 12^th^, 2020. The objective was to use the accumulative mortality rate up to that moment and a dynamic measure on how that rate is increasing or decreasing in each of the selected countries. This time span is critical when comparing the six (6) countries to determine the overall rate of mortality, and how the rate of mortality has decreased or increased in small units of time to accurately measure the prevalence. The three (3) day time periods were selected to account for the lags in reporting deaths in each of the countries, particularly with determining the cause of death as COVID-19 and the different methods of reporting data in each country. Therefore, the three (3) day lag is intended to account for these differences and standardize a time period in the series. The final variable to add a predictive deceleration variable is the moving average since peak increase, the post peak increase of mortality rate (MAPOST INC). This variable can determine the sustainability of the deceleration of COVID-19 in the six (6) selected countries.

This model was constructed as a comparative analysis with several measures. The first was the number of days from the first reported COVID-19 case to peak increase in mortality rate (PINC MR). This measure is important for several reasons. The first is early recognition of a new infectious disease and how and when the infectious disease is discovered, reported, and addressed by authorities. Another important measure used is the number of days between day of first reported death and PINC MR. This measure, and these two measures taken together can formulate the speed, intensity, prevalence, and lethality of the infectious disease in a given country.

Other measures in the analyses included the number of days between the date the COVID-19 was first reported in each of the six (6) countries and peak increase in mortality rate (PINC MR) and the number of days between the date of the first reported death in each of the six (6) countries and PINC MR. These measures were added to determine the speed and intensity of the spread of COVID-19 and to compare if there were significant differences in the expedience of first reporting COVID-19 in the six (6) selected countries. The objective of this exploratory analysis is to create and apply a time-series model to compare the time dimension of how the initial outbreak of COVID-19 within the six (6) selected countries’ mortality rates began to show signs of slowing and/or decelerating with the purpose to provide another analytical tool to assist policy decisions.

These measures are particularly important for policy makers in determining emergency response procedures such as shelter in place or suspending mass gatherings and public transportation services. The last time series measure used to regress the IMAM is the rate of mortality MA POSTINC since the day of the peak increase in mortality rate PINC MR. Once the day of PINC MR is determined based on a lower rate of mortality in the next time interval, what is the rate of mortality and how much is this rate of mortality decreasing. The model is designed to establish a measure in that consecutive days or weeks of peak increase can signal a significant deceleration of an infectious disease.

**Table One.** Number of Days from First Reported Case and Death and Date of Peak Increase

| <u>Country</u> | First Reported COVID-19 | First Reported COVID-19 | Date of Peak Increase Change | Number of Days from first Reported Case to Peak Increase Change | Number of Days from first Reported Death to Peak Increase Change |
| --- | --- | --- | --- | --- | --- |
|  | Case | Death |  |  |  |
| China | 12/8/2019 | 1/21/2020 | 4/17/2020 | 131 | 87 |
| Italy | 1/31/2020 | 2/23/2020 | 3/30/2020 | 59 | 36 |
| France | 1/27/2020 | 2/16/2020 | 4/8/2020 | 72 | 52 |
| Germany | 1/24/2020 | 3/10/2020 | 4/17/2020 | 84 | 38 |
| Spain | 2/10/2020 | 3/5/2020 | 3/30/2020 | 49 | 25 |
| UK | 1/31/2020 | 3/1/2020 | 4/29/2020 | 97 | 60 |
| US | 1/22/2020 | 3/3/2020 | 5/2/2020 | 109 | 60 |
|  |  |  | **Mean Days** | **86** | **51** |

## Results

The first day COVID-19 case was reported in the Index Country (China) was December 8, 2019. The next five (5) countries in chronological order are the United States (January 22, 2020), Germany (January 24, 2020), France (January 27, 2020), Italy (January 31, 2020), United Kingdom (January 31, 2020) and Spain (February 2, 2020). The first reported COVID-19 death in the Index Country (China) was January 21, 2020 followed by France (February 16, 2020), Italy (February 23, 2020), United Kingdom (March 1, 2020), The Moving Average change was then calculated in the sixteen (16) time periods. The purposed to determine if there is sustained deceleration in the change of rate of mortality. The model is designed to have the ability to predict that a country must sustain three consecutive time periods of deceleration to consider a decrease in COVID-19.

The Mean number of days from the first reported COVID-19 case from the day of peak increase in mortality rate (PINC MR) was eighty-six (86) days. The number of days ranged from forty-nine (49) (Spain) to 131 (China). The Mean number of days from the first reported COVID-19 case from the day of the first reported death to PINC MR was fifty-one (51) days ranging from twenty-five (25) days (Spain) to eighty-seven (87) days (China).

**Table Two.**
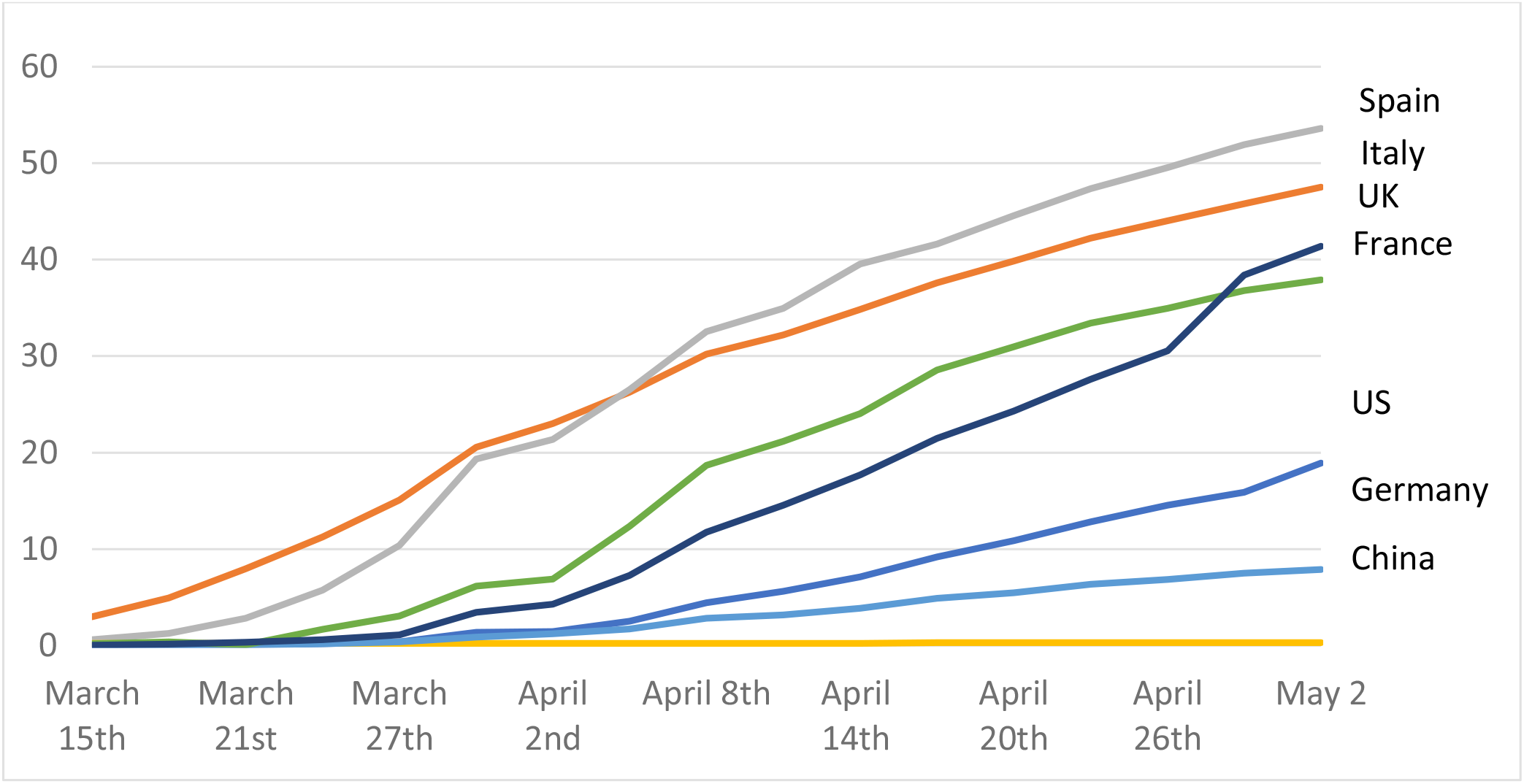
Rate of Mortality in Six (6) Selected Countries from March 15, 2020 to May 2, 2020

### Peak Increase Change in Mortality Rate (PINC MR) and Moving Average Post Increase (MA POSTINC) and evidence of Consistent Deceleration Model (CDM)

The most critical variables in the time series design was the day of peak increase in change of mortality rate (PINC MR). The earliest PINC MR was experienced by Spain (25 days from first reported death) and Italy (36 days from first reported death) on March 30^th^, 2020 in that Spain’s mortality rate peaked at 19.3 and the PINC MR was 9.0. Italy’s mortality rate on March 30^th^, 2020 was 20.5 and the PINC MR was 5.4. By contrast, the PINC MR for China occurred on April 17^th^, 2020 (119 days after the first reported death), the United Kingdom was on April 29^th^, 2020, (97 days from first reported death) and the PINC MR was not experienced in the United States until May 2^nd^, 2020, 109 days after the first reported death. France (April 8^th^, 2020 PINC MR, 72 days after first reported death) and Germany (PINC MR, April 17^th^, 2020, 84 days after first reported death) were more aligned around the average days from first reported death and PINC MR.

The overall of the objective was to create a tool to quantitatively predict deceleration of reported cases of the virus based on a Moving Average since Peak Increase (MAPINC). The MAPINC is the moving average of the reported deaths since the PINC MR. The Consistent Deceleration Model (CDM) accounts for the PINC MR and the MAPINC and the MAPINC was remain below the PINC MR for an extended period of time to display evidence that the virus is slowing. For example, policy makers should adopt a decision that included at least four (4) or more time periods (in the case of this analysis, 12 days) to determine if the virus has displayed significant signs of deceleration. Based on the CDM used in this study period, Spain’s MAPINC was 3.1 from March 30^th^, 2020 to May 2^nd^, 2020 and experienced four time periods of a moving average death rate (MADR) on April 18^th^, 2020 and remained under 3.1 until May 2^nd^. 2020. Italy had a MPINC of 2.4 and on April 24^th^, 2020 started a four-time period CDM. As of May 2^nd^, 2020, the United States and United Kingdom did not show signs of consistent slowing based on the Consistent Deceleration Model (CDM).

**Table Three.**
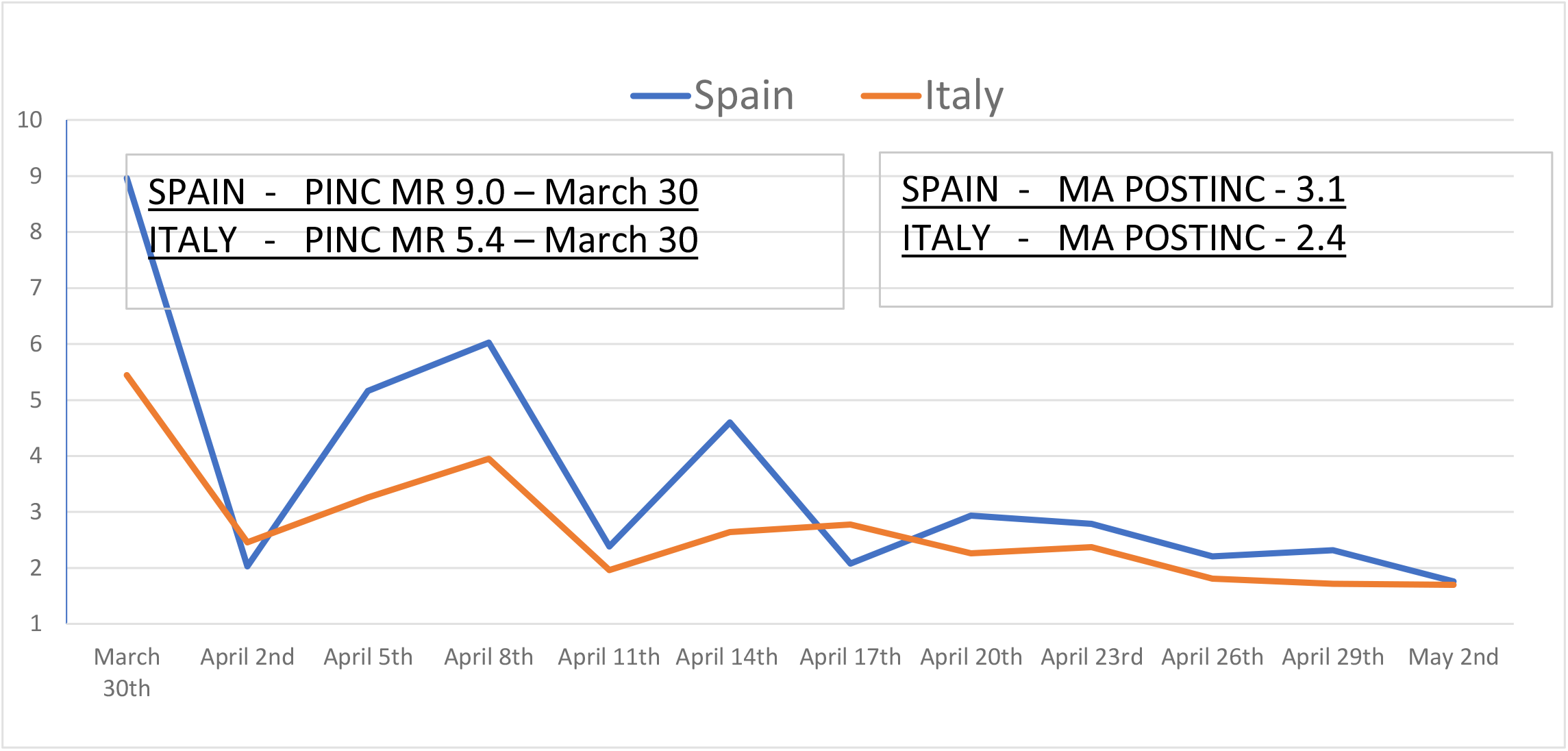
Spain and Italy PINC MR and MA POST INC

**Table Four.**
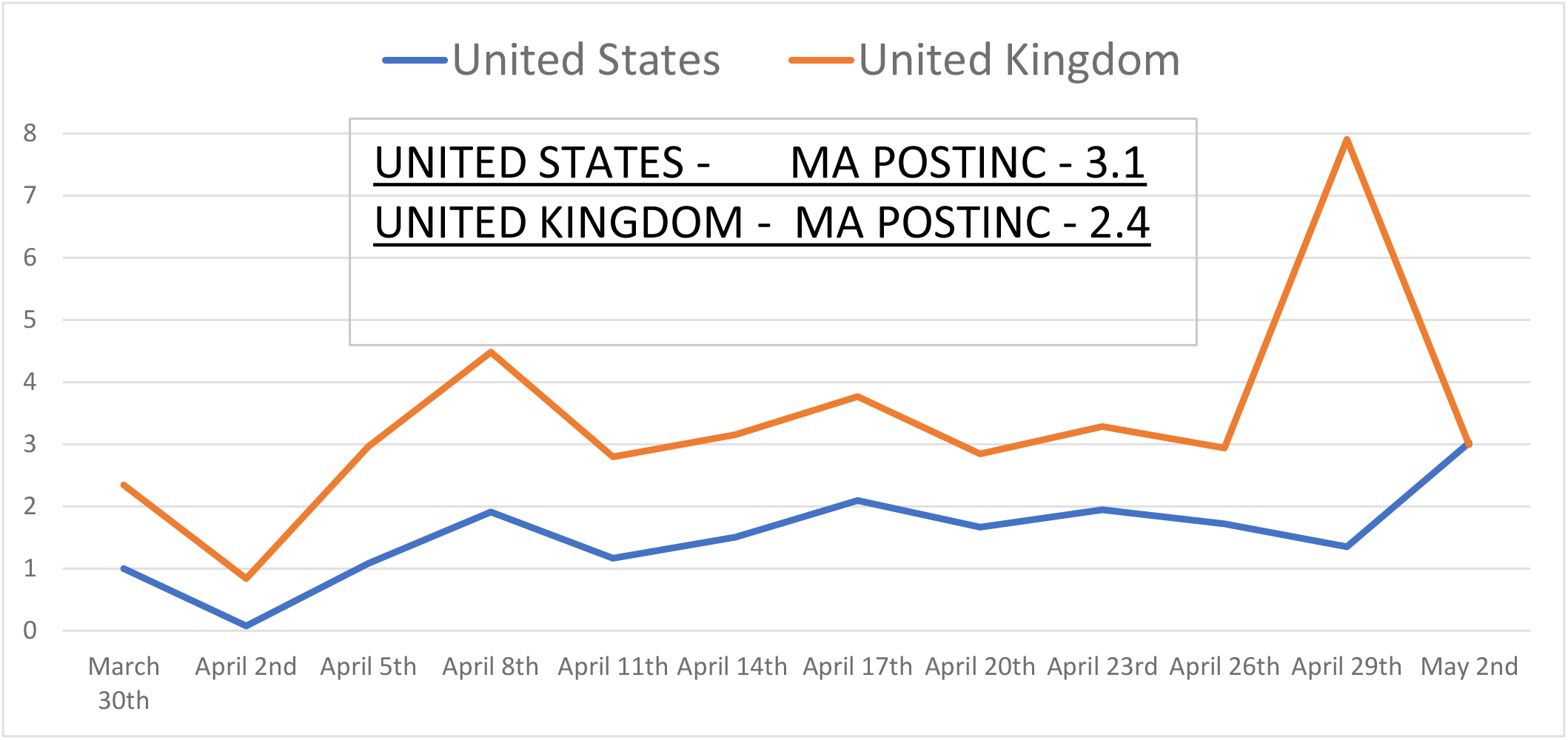
United States and United Kingdom Moving Average Post Increase Mortality Rate

## Discussion

The Consistent Deceleration Model (CDM) is a work in progress using several moving average models with dynamic and static data. Ideally, the CDM is also designed to be flexible for each country and individual regions within countries where an outbreak is most susceptible. The CDM is designed to predict consistent sustainable COVID-19 first starts to decelerate and sustain deceleration based on individual occurrences and outbreaks in each country. This is particularly important to determine given the intensive spread of the COVID-19 virus.

Additionally, this model was constructed to compare countries that were severely affected to determine how the rate of acceleration and deceleration of the mortality rates varied by country. This model should be utilized as one of many tools to determine the intensity and prevalence of the outbreak and when there are quantifiable signs of a significant deceleration.

## Data Availability

I have the data files saved and they are available for anyone to view or analyze upon request.

